# Adaptation needs of an intervention to facilitate tobacco cessation among men attending emergency departments in Nairobi, Kenya

**DOI:** 10.64898/2026.09.02.26361107

**Authors:** Mari Armstrong-Hough, Thomas Kedera, Miriam Bosire, Melvine Obuya, Grace Soma, Jessica Rayo, Lorien Abroms, Christine Ngaruiya

**Affiliations:** New York University School of Global Public Health, Department of Epidemiology and Department of Social & Behavioral Sciences; Stanford University School of Medicine

## Abstract

**Background:** Three-quarters of cancer deaths globally are in low- or middle-income countries. Cost-effective tobacco cessation interventions are critical to reducing cancer burden in these settings. Interventions that reach men, who are more likely to smoke and less likely to be engaged in primary care, are particularly important in Kenya, where 84% of smokers are men. Engaging men using a text-based mHealth program is a promising strategy to promote cessation. Text-based quit counseling using Text2Quit, a US-based mHealth intervention, has been demonstrated to be effective in high-income settings. We aimed to identify barriers to using Text2Quit among Kenyan men who smoke and optimize Text2Quit for Kenya.

**Methods:** We invited men who smoke in Nairobi, Kenya to participate in intercept interviews using a Text2Quit prototype adapted for Kenya. A multinational team of tobacco experts, designers, Kenyan physicians, and implementation researchers carried out a rapid analysis of interview transcripts to identify barriers to its usability, relatability, and appeal. These barriers and proposed solutions were enumerated and ranked using nominal group technique with rank-order voting.

**Results:** Thirteen men participated in interviews using a Text2Quit-Kenya prototype. Participants identified rigid response formats as a barrier to its usability, long text messages as a barrier to relatability, and confusion about cost as a barrier to its appeal. Twenty-six proposed solutions were identified by the nominal group technique process and five were prioritized based on importance to stakeholders and feasibility: 1) enable minor deviations in patient responses, 2) reduce length of messages, 3) clearly communicate that Text2Quit is free of charge, 4) provide brief in-person counseling at enrollment, including training on how to find and reply to texts and setting a quit date before texts begin, and 5) provide individualized follow up with users.

**Conclusions:** Barriers to uptake of Text2Quit among Kenyan men who smoke are addressable through a systematic adaptation process.

## Introduction

Low- and middle-income countries account for three-quarters of all cancer deaths. Cost-effective tobacco cessation interventions are critical to reducing cancer burden in these settings (Abroms et al., 2014; Reisinger et al., 2019). In Kenya, according to the 2015 WHO STEPS national survey, around one in eight people (13.5%) reported being current tobacco users. Most active tobacco users were male (n = 507/605, 83.8%) and younger than 50 years old (n = 468/605, 77.4%), with the average start age being 21 (20.6, 95% CI 19.3-21.8). Men had nearly seven times higher odds of being tobacco users compared to women (OR 7.63, 95% CI 5.63-10.33). They also constitute 84% of smokers in Kenya. Therefore, interventions that reach men, who are more likely to smoke and less likely to be engaged in primary care, are particularly important in Kenya (Ngaruiya et al., 2018).

Engaging men using a text-based mobile health (mHealth) program is a promising strategy to promote tobacco cessation. mHealth applications like telephone-based short message service (SMS) text messaging have been shown to facilitate quitting in some populations. A Cochrane Review of mobile phone text messaging and app-based interventions for smoking cessation found moderate-certainty evidence of benefit. However, settings for delivery of tobacco cessation interventions in LMICs differ markedly from the high-income settings in which most evidence-based cessation programs were originally developed and evaluated. For example, more than half of people in Kenya use feature phones (Communications Authority of Kenya, 2025). mHealth interventions to promote tobacco cessation cannot be adopted wholesale from high-income settings; they must be adapted to meet the needs and preferences of Kenyan users.

Text2Quit is an interactive mobile health program designed to help individuals quit smoking by providing personalized support through text messages, emails, and a dedicated website (Abroms et al., 2014). The mobile health program delivers automated text messages timed around the user’s selected quit date, offering advice, encouragement, reminders, and prompts to help track smoking behaviors and cravings (Abroms et al., 2014). Messages are tailored based on different user characteristics (i.e., name, quit date, and motivations for quitting) and are informed by social cognitive theory and U.S. clinical practice guidelines for tobacco cessation (Abroms et al., 2014). Participants can also request additional support by sending specific keywords or by accessing a personalized website and supportive emails (Abroms et al., 2014). In a randomized controlled trial conducted in the U.S., Text2Quit significantly enhanced both biochemically confirmed and self-reported quit rates compared to control interventions, with positive results observed across different demographic groups (Abroms et al., 2014).

Text-based quit counseling using Text2Quit has been demonstrated to be effective in high-income settings. However, this evidence-based intervention has not previously been adapted, implemented, or evaluated in a LMIC setting. We aimed to identify barriers to using Text2Quit among Kenyan men who smoke and optimize Text2Quit for Kenya using a systematic adaptation process based on intercept interviews with users, stakeholder participation, and nominal group technique.

## Methods

We designed a qualitative user experience study to adapt Text2Quit to the needs, preferences, and perceptions of men who smoke in Kenya.

First, a multinational team including two Kenyan physicians preliminarily adapted the content of Text2Quit to the cultural and linguistic setting, including adapting the modality of intervention delivery to text message alone. We then invited men who smoke who presented in the emergency department of Kenyatta National Hospital (KNH) in Nairobi, Kenya to participate in intercept interviews using the preliminarily adapted Text2Quit prototype. KNH is the largest referral and teaching hospital in East Africa, serving approximately 3 million people from Kenya and neighboring countries (Kenyatta National Hospital, 2024). Eligible patients were invited to individually engage with the Text2Quit prototype in a private consultation room. A trained research assistant carried out an intercept interview as the participant engaged with the prototype from their personal mobile phone, including receiving and sending messages. A second trained research assistant served as a notetaker, recording the participants’ use of and comfort with the Text2Quit prototype. Using a cognitive interview format, participants were then probed regarding their understanding of the tool, perceptions of the user interface, and expectations for its operation, including frequency and timing (Sexton et al., 2023). Interviews were transcribed for analysis. Research assistants also produced structured debriefing reports summarizing participants’ responses.

A multinational team of tobacco experts, designers, Kenyan physicians, and implementation researchers then carried out a rapid analysis of interview transcripts to identify barriers to the usability, relatability, and appeal of the Text2Quit prototype. Using a novel participatory strategy based on nominal group technique (NGT) (Dunham, 1998; Centers for Disease Control and Prevention [CDC], 2018), the adaptation team read interview transcripts and debriefing reports and prepared worksheets (Supplement S1) to nominate candidate adaptations based on test user experiences. The adaptation team enumerated and ranked barriers and proposed solutions using the nominal group technique (NGT) with rank-order voting (Dunham, 1998; CDC, 2018). This process ensures transparent, democratic decisions that reflect diverse expertise and experience.

NGT is a formal process to generate ideas and reach decisions through small-group discussion (Dunham, 1998; CDC, 2018). The process is designed to facilitate equal contribution from all team members and to maximize the efficiency of the discussion. A study team member with experience facilitating design research using NGT (MAH) facilitated the process in five steps:

1. *Generate ideas*: In the first step, all participants independently reviewed the transcripts and wrote down ideas for adaptation in response to user experiences in brief phrases or statements using a worksheet to focus responses on improving the usability, relatability, and appeal of the evidence-based intervention.
2. *Record ideas*: Participants engaged in a feedback session to briefly present the ideas that each team member contributed. Completed worksheets were used to ensure that all team members’ ideas were put forward.
3. *Discuss ideas*: Using a round robin format, participants discussed every submitted idea to ensure that it was well understood. For each idea, the facilitator asked, “Are there any questions or comments group members would like to make about the item?” The objective of this step was to understand “the logic and the relative importance” of each suggestion.
4. *Prioritize ideas*: Finally, participants voted on the priority of the suggested ideas. Team members each privately chose the five ideas for intervention adaptation that they thought to be the highest priority, then rank-ordered those ideas.
5. *Vote*: The group submitted their rank-order votes to an outside study coordinator, who tabulated and announced the results.

Following the vote, the five top-ranking adaptations were integrated into the mature prototype for a planned pilot trial.

## Results

Thirteen men who smoke participated in interviews using a Text2Quit-Kenya prototype (**Table 1**). Participants ranged in age from 36 to 67 years of age. More than half (n=7, 54%) were employed and reported a median annual individual income of 109,500 Shillings (range: 37,800 - 219,000 Shillings). Participants had presented to the emergency department with a range of complaints: respiratory symptoms, injury, chest pains, cancer complications, and toothache.

**Table 1.**
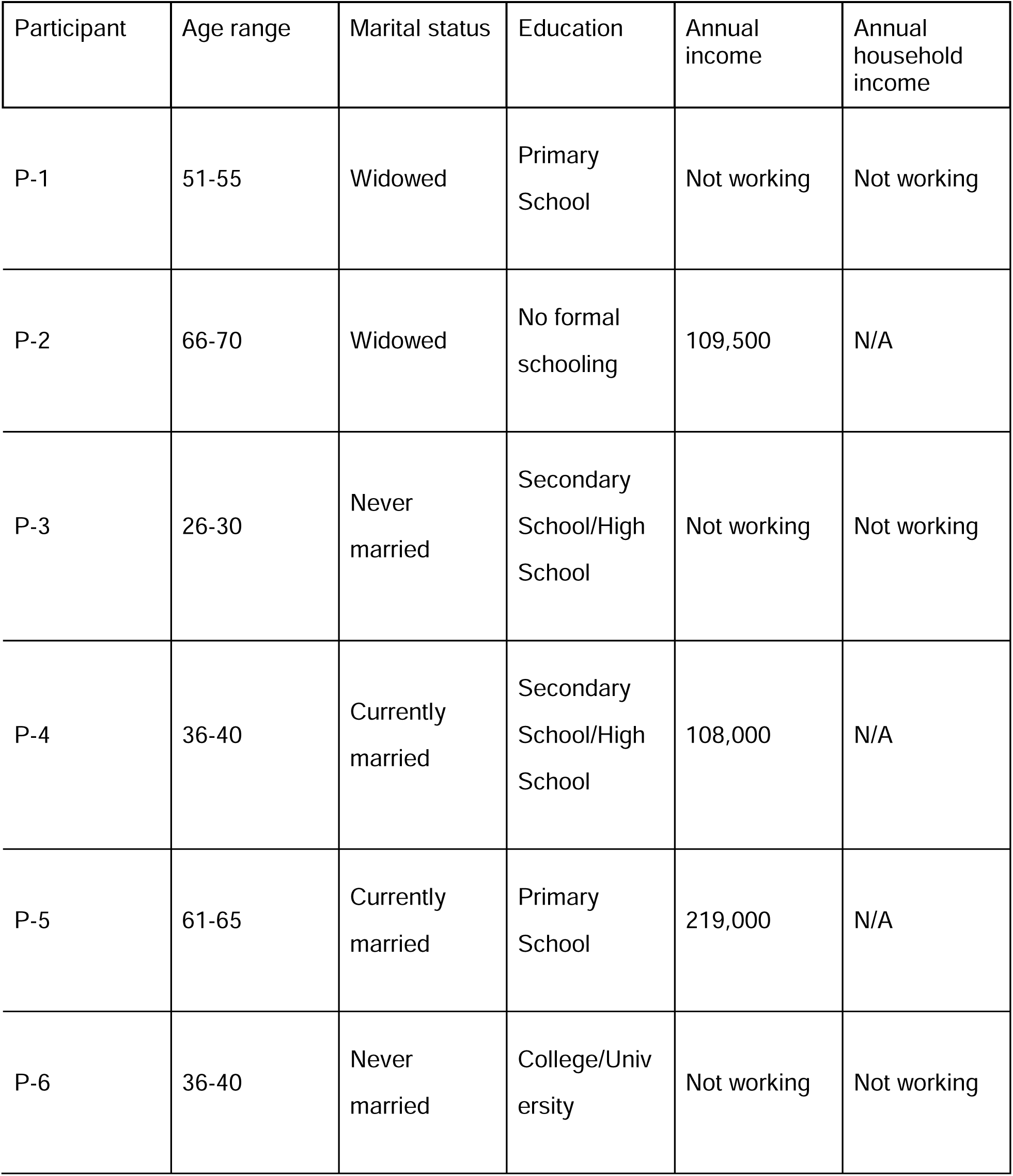
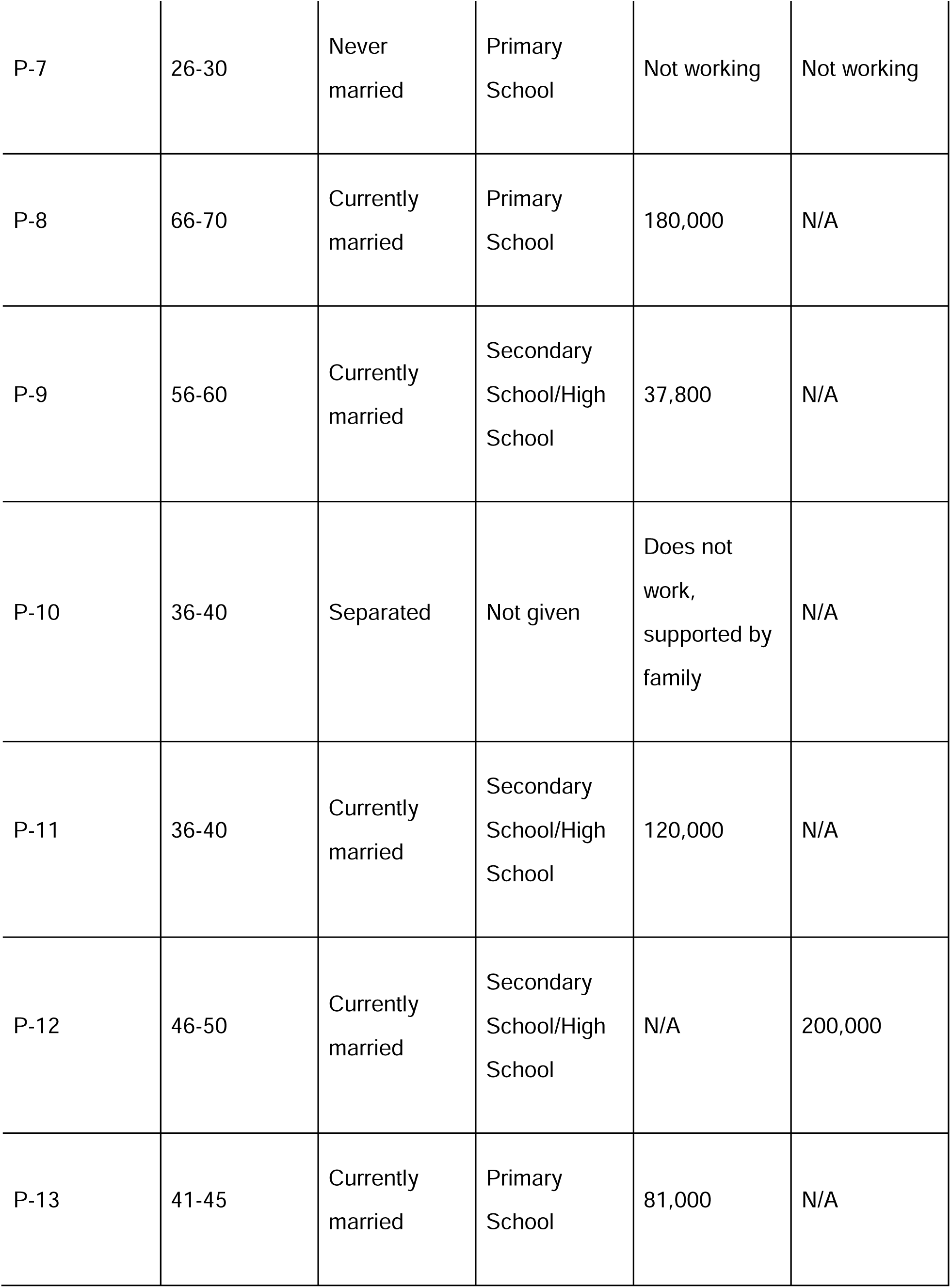
Description of the sample.

| Participant | Age range | Marital status | Education | Annual income | Annual household income |
| --- | --- | --- | --- | --- | --- |
| P-1 | 51-55 | Widowed | Primary School | Not working | Not working |
| P-2 | 66-70 | Widowed | No formal schooling | 109,500 | N/A |
| P-3 | 26-30 | Never married | Secondary School/High School | Not working | Not working |
| P-4 | 36-40 | Currently married | Secondary School/High School | 108,000 | N/A |
| P-5 | 61-65 | Currently married | Primary School | 219,000 | N/A |
| P-6 | 36-40 | Never married | College/University | Not working | Not working |
| P-7 | 26-30 | Never married | Primary School | Not working | Not working |
| P-8 | 66-70 | Currently married | Primary School | 180,000 | N/A |
| P-9 | 56-60 | Currently married | Secondary School/High School | 37,800 | N/A |
| P-10 | 36-40 | Separated | Not given | Does not work, supported by family | N/A |
| P-11 | 36-40 | Currently married | Secondary School/High School | 120,000 | N/A |
| P-12 | 46-50 | Currently married | Secondary School/High School | N/A | 200,000 |
| P-13 | 41-45 | Currently married | Primary School | 81,000 | N/A |

In intercept interviews, participants identified rigid response formats as a barrier to the usability of Text2Quit in Kenya, long text messages as a barrier to relatability, and confusion about cost of using Text2Quit as a barrier to its appeal.

### Usability

Several participants struggled with the rigid response formats required to interact with Text2Quit by text. For example, one respondent responded to a test message with a date in an unaccepted format: “[DD]th [MM]/[YYYY]” (P-12). He subsequently received a message informing him that the message was in an invalid format. He expressed frustration but persisted, again entering “[DD]^th^ [MM]/[YYYY]” and resending the message. When he again received a message informing him that his message was in an invalid format, he gave up.

### Relatability

Many participants also struggled to relate to the long text messages. Although the text had already been preliminarily adapted to the Kenyan context and translated to Swahili, most participants were not accustomed to reading long messages on their mobile phone. This issue was exacerbated by the use of feature phones, which required participants to scroll down to finish reading longer messages. One participant described reading the messages as “a torture to my brain” (P-5). Another did not respond to the text messages he received and read during the intercept interview. He explained his unresponsiveness to the research staff, saying, “there is something I don’t understand.” (P-4)

### Appeal

Many participants remained unsure about the cost of participating in Text2Quit, even after the introductory message and counseling indicating that there was no cost to use the program. For example, one participant (P-6) said he did not respond to any of the Text2Quit messages because he “did not have any airtime.” Participants worried they could be charged for use and reported that this reduced the appeal of using it regularly.

#### Adaptations

Seven study team members participated in the nominal group technique process to generate adaptations in response to these barriers, including two Kenyan research assistants, one Kenyan physician, one Kenyan-American physician, one Kenyan developer, one non-Kenyan social scientist, and the original non-Kenyan creator of Text2Quit. Sixteen proposed solutions were identified during the rapid analysis and an additional ten solutions identified during the discussion (**Table 2**). Following rank-order voting, five were prioritized based on importance to stakeholders and feasibility: 1) enable minor deviations in patient responses, 2) reduce length of messages, 3) clearly communicate that Text2Quit is free of charge, 4) provide brief in-person counseling at enrollment, and 5) provide individualized follow up with users.

**Table 2.**
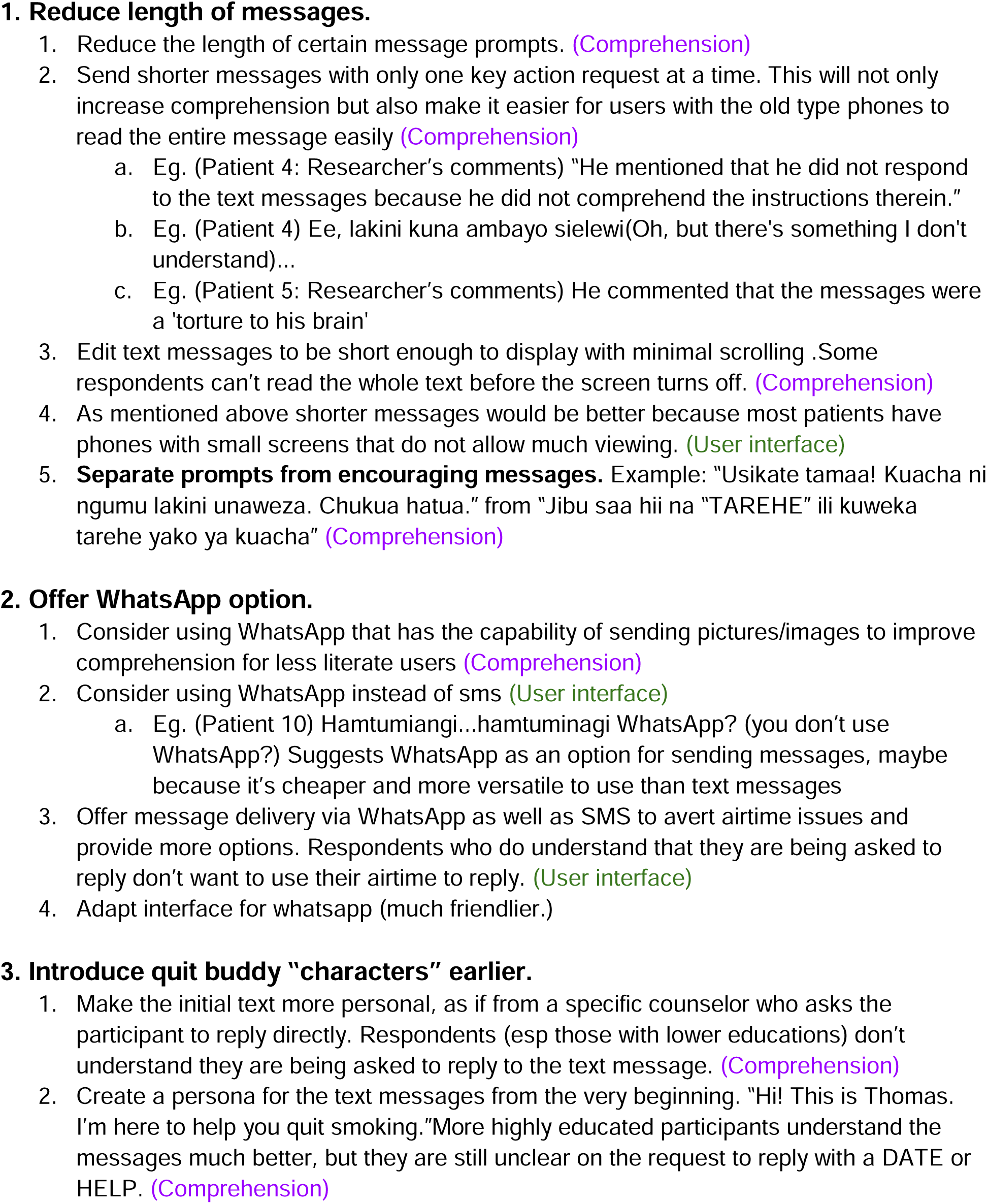
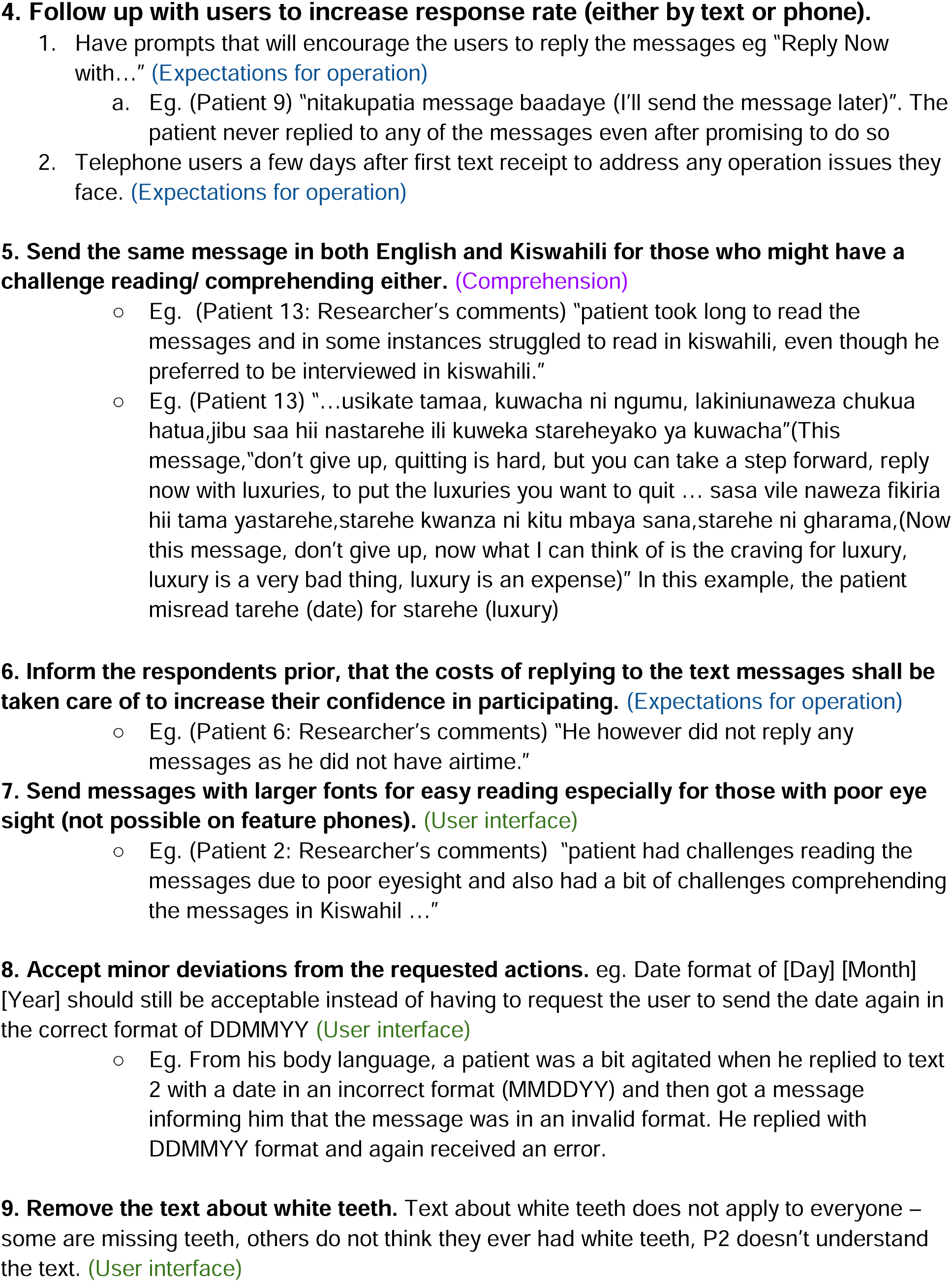
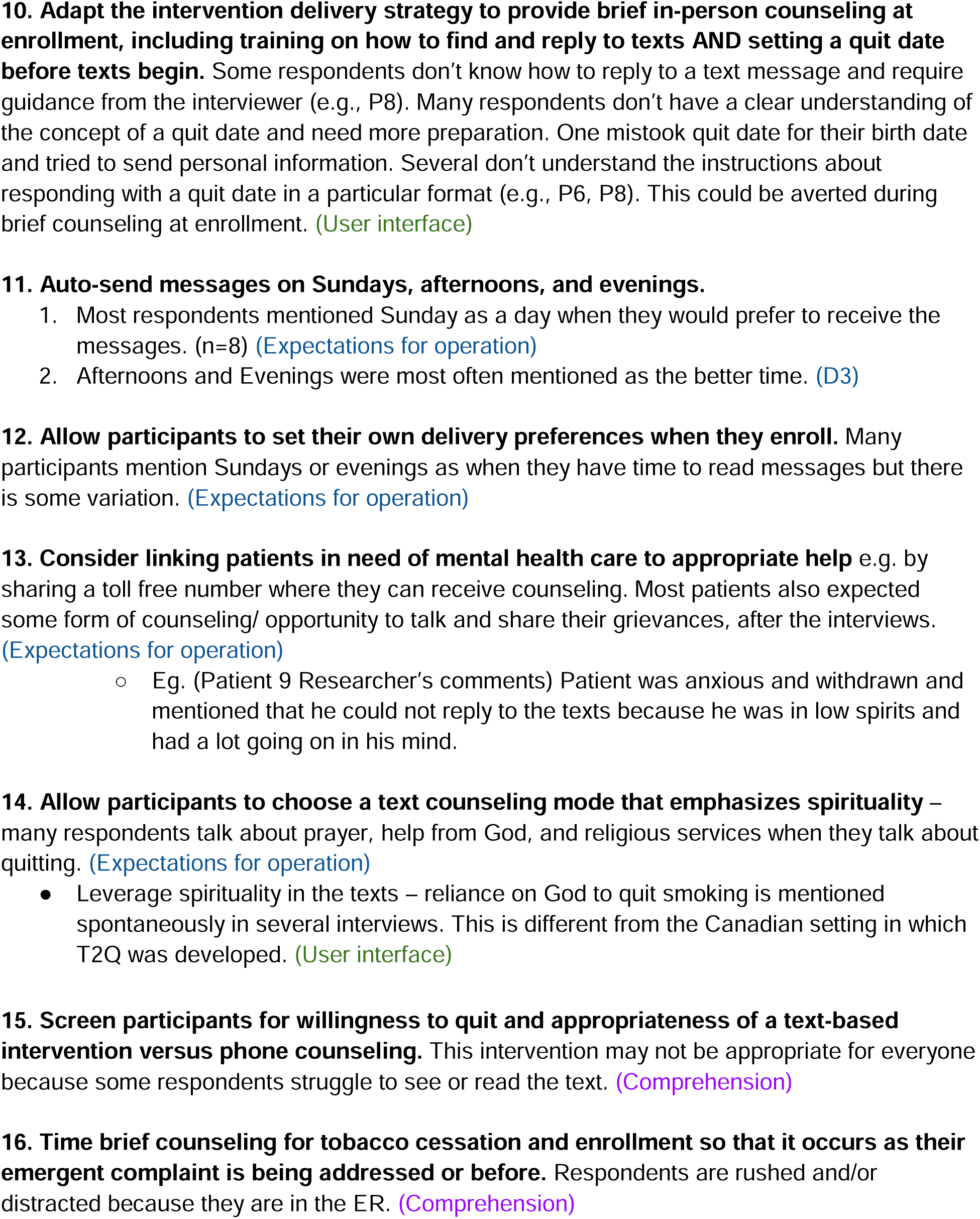
Adaptation candidates generated by nominal group technique.

**Table 3.** Nominal group technique rank-order voting results.

| Rank | Adaptation candidate | Votes | Disposition |
| --- | --- | --- | --- |
| 1 | **Allow minor deviations in participant responses (e.g., alternative date formats, common misspellings, and shorthand)** | 16 | **Adopted** |
| 2 | **Reduce message length and limit each message to one key action** | 15 | **Adopted** |
| 3 | **Clearly communicate that participation and replies are free of charge** | 10 | **Adopted** |
| 4 | **Provide brief in-person counseling at enrollment, including quit-date setting and text-reply training** | 9 | **Adopted** |
| 5 | **Add follow-up prompts or calls to increase response rates** | 8 | **Prioritized for future implementation** |
| 6 | Let users choose their preferred language rather than sending bilingual messages | 7 | Not adopted |
| 7 | Send messages in both English and Kiswahili | 5 | Not adopted |
| 8 | Link users with mental health support when needed | 5 | Not adopted |
| 9 | Offer WhatsApp as an additional delivery option | 4 | Not adopted |
| 10 | Allow users to set preferred delivery times at enrollment | 4 | Not adopted |
| 11 | Provide numbered response options | 3 | Not adopted |
| 12 | Clarify the program name because “Text2Quit” loses meaning in Kiswahili | 2 | Not adopted |
| 13 | Remove the message about “white teeth” | 1 | Not adopted |
| 14 | Schedule automated messages on Sundays, afternoons, and evenings | 1 | Not adopted |

*Note. Items prioritized through NGT voting are bolded and shaded.*

### Minor deviations in response

The highest-scoring adaptation was to expand the range of deviations allowed in responses. For example, rather than accepting dates in only one format (DDMMYY), date formats in other formats should be accepted and subsequently reconfirmed with the user to prevent misunderstanding. Similarly, shorthand and common misspellings should be added to the acceptable responses (e.g., “Y”, “ys”, “ye” would all be accepted as indicating “yes”).

### Length of messages

The second highest-scoring adaptation was to reduce the length of messages. In discussion, the panel identified four design principles to improve the readability of messages:

1. Send messages with only one key action request at a time.
2. Edit messages to enable display with minimal scrolling on older model feature phones.
3. Prioritize shorter messages and break up longer messages when possible.
4. Separate prompts from encouraging messages.

### Reiteration that Text2Quit is free of charge

Panel members voted to adapt the delivery of Text2Quit to repeatedly reinforce that it is free of charge to users and, specifically, that the texts sent to participants would not deplete users’ prepaid cellular service accounts. This message will be reinforced at initiation and through periodic reminder messages.

### In-person counseling at enrollment

Panel members also voted to adapt the intervention delivery strategy to provide brief in-person counseling at enrollment. During discussion, the panel agreed this should include two components: setting a quit date before texts begin and training users on how to find and reply to texts on the participant’s own device during enrollment.

### Individualized follow-up

Finally, panel members voted to adapt the intervention to include more individualized follow-up. Specifically, the panel discussed two ways to improve follow-up in order to increase engagement with the intervention: 1) automatically generated prompts encouraging users to reply after one day of non-response and 2) a personalized telephone call from a Text2Quit counselor a few days after first text receipt to address operational issues they may face as they begin the program.

## Discussion

In this intercept interview-based, participatory analysis of barriers to using Text2Quit among Kenyan men who smoke, we found that participants struggled with rigid response formats, skipped reading long text messages, and lost enthusiasm when they misunderstood the free-of-charge program to require them to pay for text messages or airtime. Through a participatory process using nominal group technique, we identified and prioritized key needs for adapting Text2Quit for Kenya: minor deviations in patient responses, shorter messages, clear communication that Text2Quit is free of charge, brief in-person counseling at enrollment, and individualized follow-up with users.

Notably, the intercept interview format made it clear that many participants did not finish reading long text messages. One reason for this may be that most participants used feature phones rather than smartphones. Feature phones–mobile phones that offer only core functions such as voice calls, SMS, a physical keypad, and a non-touchscreen display–remain common in Kenya and in many parts of Africa. Screens on feature phones are small and reading a longer text message requires the user to scroll down, which may deter users from finishing longer messages. However, it is important to recognize that skipping long messages may simply be a revealed preference among potential Text2Quit users for simpler and more straightforward messages. A previous evaluation of an mHealth evaluation in rural Kenya to prevent mother-to-child-transmission of HIV noted that the 160-character SMS limit can be helpful because it forces messages to be simple and understandable for low-literacy users (Nordberg et al., 2024).

Most research in Kenya and other LMICs evaluates whether SMS-based mHealth interventions are effective rather than how characteristics such as length may impact uptake and effectiveness. Our finding that some participants stop reading after just a few lines of text suggests that future research should extend beyond indirectly addressing length via literacy and content simplicity. Of note, the patient population in this study reflects health literacy realities seen commonly in public-sector healthcare settings in Africa. Findings from this population may offer important insights into the equitable implementation and scalability of digital health interventions in resource-variable contexts, with particular attention to even more marginalized populations such as those residing (or receiving care) in informal settlements, a burgeoning population (Harsch et al., 2025).

Intercept interview responses also pointed to another barrier reducing engagement with Text2Quit messages: the perception that responding to text messages would incur charges. Although participants received both written and oral explanations that text message engagement with Text2Quit would be free of charge, multiple participants hesitated to respond because of cost. This is consistent with evaluations of other mHealth interventions in the region. In an analysis of program data from Mobile for Reproductive Health (m4RH), a national SMS-based health communication service in Tanzania, less than half of users were willing to pay for SMS service; authors argued that SMS charges could deter use among the poorest and most vulnerable users (Mangone et al., 2016). In Kenya and in many LMIC, mobile phone users often pay per message. Our findings suggest that even when the program is designed not to incur charges for participants, users may assume that they will bear costs. It was also evident that some education or sensitization on the tool, including assuaging fears on costs, demonstrating usability, login and troubleshooting were key. This would also be helpful in the case of similar intervention implementation in comparable settings.

Finally, the nominal group technique process led to the prioritization of five key adaptations. These adaptations were directly incorporated into an ongoing trial testing the adapted Text2Quit intervention against a control arm receiving standard of care, where implementation and clinical outcomes will be assessed. The latter will be indicated by tobacco cessation, carbon monoxide testing, and self-report.

## Conclusion

Cost-effective tobacco cessation interventions are critical to reducing morbidity and mortality in LMICs. Interventions that reach men are particularly important in Kenya, where 84% of smokers are men. SMS-based tobacco cessation interventions are low-cost, reliable, and widely accessible to men who smoke. We systematically adapted Text2Quit, an evidence-based mobile phone intervention originally generated for a general population in a high-income setting, for men who smoke in Kenya. We found that barriers to uptake of Text2Quit among Kenyan men who smoke were addressable through a systematic adaptation process that engaged both end-users and implementers. NGT remains an effective approach for extracting input and consolidating suggestions, in an equitable and inclusive fashion, for adaptation of technical interventions in LMICs as in the current study.

## Data Availability

All data produced in the present study are available upon reasonable request to the authors

## References

Abroms LC, Boal AL, Simmens SJ, Mendel JA, Windsor RA. A randomized trial of Text2Quit: a text messaging program for smoking cessation. Am J Prev Med. 2014;47(3):242–250. doi:10.1016/j.amepre.2014.04.010

Ngaruiya C, Abubakar H, Kiptui D, Kendagor A, Ntakuka MW, Nyakundi P, et al. Tobacco use and its determinants in the 2015 Kenya WHO STEPS survey. BMC Public Health. 2018;18(Suppl 3):1223. doi:10.1186/s12889-018-6058-5

Reisinger SA, Thomson N, Gunukula S, Ahmad I, Spigner C. Cost-effectiveness of community-based tobacco dependence treatment interventions: Initial findings of a systematic review. Prev Chronic Dis. 2019;16:E162. Published 2019 Dec 12. doi:10.5888/pcd16.190232

Kenyatta National Hospital. Our History. Kenyatta National Hospital website. https://knh.or.ke/index.php/history/. Published August 21, 2024. Accessed October 17, 2025.

Communications Authority of Kenya. Mobile, data, and digital services on the rise, CA report shows [Internet]. Nairobi: Communications Authority of Kenya; 2025 Jun 30 [cited 2026 May 21]. Available from: https://www.ca.go.ke/mobile-data-and-digital-services-rise-ca-report-shows

Sexton O, Pilley S, d’Ardenne J, Bull R. Cognitive interviewing and what it can be used for [Internet]. National Centre for Research Methods; 2023 [cited 2026 May 21]. Available from: https://www.ncrm.ac.uk/resources/online/all/?id=20816

Dunham RB. Nominal group technique: a users’ guide [Internet]. Madison (WI): University of Wisconsin; 1998 [cited 2026 May 21]. Available from: https://sswm.info/sites/default/files/reference_attachments/DUNHAM%201998%20Nominal%20Group%20Technique%20-%20A%20Users%27%20Guide.pdf

Centers for Disease Control and Prevention [CDC]. Gaining consensus among stakeholders through the nominal group technique. Evaluation Briefs No. 7 [Internet]. Atlanta (GA): Centers for Disease Control and Prevention; 2018 Aug [cited 2026 May 21]. Available from: https://www.cdc.gov/healthy-youth/php/program-evaluation/pdf/brief7.pdf

Nordberg B, Kaguiri E, Chamorro de Angeles KJ, Gabriel EE, van der Kop ML, Mwangi W, et al. The use, adherence, and evaluation of interactive text-messaging among women admitted to prevention of mother-to-child transmission of HIV care in Kenya (WelTel PMTCT). BMC Pregnancy Childbirth. 2024;24:25. doi:10.1186/s12884-023-06194-0

Harsch S, Weber LS, Kassaman D, Kailemia PN, Oria VO. Cancer health literacy in Kenya: a scoping review on evidence, concept and a situational analysis of interventions. Front Public Health. 2025;13:1527400. doi:10.3389/fpubh.2025.1527400

Mangone ER, Agarwal S, L’Engle K, Lasway C, Zan T, van Beijma H, et al. Sustainable cost models for mHealth at scale: modeling program data from m4RH Tanzania. PLoS One. 2016;11(1):e0148011. doi:10.1371/journal.pone.0148011

